# Managing uncertainty and complexity during a public health emergency: Understanding the immediate and ongoing effects of the COVID-19 epidemic on a global Neglected Tropical Disease program

**DOI:** 10.1101/2024.10.25.24316139

**Authors:** E. Sutherland, R. Stelmach, N. Warren, J. Jackson, B. Allen, U. Mwingira, M. Brady, J. Ngondi, L. Hernandez, G. Dahal, G. Kabona, M. Telfort, F. Oydediran, F. Seife, H. Sitoe, M. Baker

**Affiliations:** RTI International, RTP, NC; Republic of the Philippines Department of Health, Manila; Government of Nepal Ministry of Health and Population, Kathmandu; United Republic of Tanzania, Ministry of Health, Dodoma; Ministere de la Santé et de la Prévention: Ministere de la Sante et de l’Action Sociale, Port-au-Prince; Nigeria Federal Ministry of Health, Abuja; Ethiopia Ministry of Health, Addis Adaba; Ministério da Saúde: Ministerio da Saude, Maputo; Georgetown University, Washington D.C

## Abstract

When COVID-19 emerged as a global pandemic, the World Health Organization (WHO) recommended a pause in the delivery of neglected tropical disease preventative chemotherapy and surveillance. The Act to End NTDs | East program (Act | East) worked with country neglected tropical disease (NTD) programs to develop, support, and implement guidelines that allowed NTD service delivery and surveillance to resume. This paper examines those adaptations that Act | East made as a program to support numerous countries, over a discrete time period, to resume NTD program operations. This paper also examined how the pause and the resumption of service delivery with new guidelines and standard operating procedures in place affected program operations. Specifically, we examine delays in scheduled mass drug administration and disease surveys, coverage achieved by resumed mass drug administration campaigns, and the impact that COVID-19 had on planning and budgeting. We review which adaptations have been retained in a post-COVID-19 landscape, and which may inform NTD and other global health programs, to better respond in future public health emergencies.

## Introduction

On March 11, 2020, the World Health Organization (WHO) declared coronavirus disease (COVID-19) a pandemic. Over the following months COVID-19 spread quickly, not only infecting individuals with the virus but also affecting people’s ability to actively access healthcare [1] On April 1, 2020, WHO recommended that community-based surveys, active case-finding activities, and mass drug administration (MDA) campaigns for NTDs be postponed until further notice because of the risk of transmitting COVID-19. Ministries of Health, along with many other public service efforts, had to rapidly suspend operations [2]. Soon afterward, it became clear that we needed to plan, not for re-start after the pandemic, but rather restarting amidst a pandemic.

This paper provides an analysis of how the U.S. Agency for International Development (USAID)’s Act to End NTDs | East program (Act | East), managed through the uncertainties of those times and the effects of its choices on the progress towards disease elimination and control. This program, even at the best of times, is large and complex. The program supports Ministry-led efforts to eliminate and control five different diseases (lymphatic filariasis, onchocerciasis, schistosomiasis, soil transmitted helminthiasis, and trachoma), with a focus on MDA and disease surveillance. It operated, at the time, in support of national NTD programs in thirteen countries in Africa (Democratic Republic of Congo, Ethiopia, Mozambique, Nigeria, Uganda, Tanzania), Asia (Bangladesh, Indonesia, Laos, Nepal, Philippines, Viet Nam) and the Americas (Haiti). RTI International leads the implementation of Act | East with a consortium of partners including The Carter Center, Fred Hollows Foundation, IMA World Health, Light for the World, Results for Development, Save the Children, Sightsavers, and WI-HER.

Each of the thirteen Act | East-supported countries experienced the onset and subsequent trajectory of COVID-19 infection and disease differently. In the first year, Europe and the Americas recorded the highest numbers of COVID-19 cases, followed by Southeast Asia. At that time, Africa reported very few cases[3]. These countries had varying degrees of access to the commodities and systems that enabled testing and reporting of cases (making it hard at times to accurately interpret the numbers), as well as other COVID prevention and treatment measures. Each country managed its own COVID-19 response, of course, while influenced by prevailing politics, available information, and accessible resources.

In line with WHO recommendations and following guidance from USAID, on April 10, 2020, RTI issued an official memo to Act | East partners and staff suspending Act | East support for all community-level activities and in-person meetings. This decision affected communities at risk for NTDs, government health ministries, employees, and volunteers in Act | East supported countries all around the world.

During the suspension period, as Act | East and its partners prepared to resume activities, many—sometimes contradictory—concerns were voiced. The health and safety of the community and of health workers took utmost importance. In addition, there was concern that the health systems would crumple under the added burden of COVID-19, exacerbated by supply shortages, health worker shortages and burnout, and limited facility capacities [4–6]. There were also concerns that backlash against COVID-19 infection prevention measures could affect implementation of other health campaigns [4]. The NTD community anticipated setbacks in progress towards NTD control and elimination and feared increased NTD morbidity and mortality [6–7]. Act | East had to consider all these concerns when, on July 27, 2020, the WHO issued further interim guidance that activities could restart following a case-by-case risk assessment [8]. Several published papers about this early implementation period describe the calculus undertaken in first restarting activities during the COVID-19 pandemic [9–14].

This paper adds further to the existing literature in a several ways. First, as well as describing how the Act | East program – a large, global, multi-disease USAID program – adjusted at the onset of COVID-19, it also assesses how that approach adapted to changing conditions during the months that followed. Secondly, it provides data on the impact that the suspension had on mass drug administration coverage and on assessment of disease prevalence surveys. Finally, it includes an analysis of how the COVID-19 pandemic affected budgeting and allocation of program resources. Based on the results, we make recommendations for long-term adaptations and provide lessons learned for recovery and mitigation of future public health emergency situations, recommendations which will contribute to overall program resilience and greater global health security.

## Methods

This research aimed to describe the adaptations made in response to COVID-19 by a large USAID program supporting the control and elimination of neglected tropical diseases, and the impact on Act | East management, budgets, expenditures, and program coverage. The main research questions and sub questions assessed in this study are summarized in table 1. To address these questions, the authors took a multi-faceted approach known as an outcome harvest evaluation. Outcome harvesting is a complexity-aware qualitative evaluation methodology that triangulates data from key informant interviews with program data and documents to identify, verify, describe, and understand outcomes. In the context of outcome harvesting, an outcome is defined as “a change in the behavior, relationships, actions, activities, policies, or practices of an individual, group, community, organization, or institution” [15].

In this study, data came from qualitative findings from program document review and key informant interviews, quantitative program data on activities supported, and results of a mixed method costing analysis. Country-specific mass drug administration and survey data are limited to eight countries that Act | East supports: Ethiopia, Nigeria, Tanzania, Mozambique, Nepal, Indonesia, Philippines, and Haiti. Data were collected and analyzed for the period between March 2020 (just before the global suspension of NTD operations) and September 2021, well after operations had resumed.

### Ethics

Because this study collated and analyzed data collected as part of ongoing monitoring of public health programs with the purpose of describing public health program implementation without the goal of generalizing this knowledge or experience to other settings, this assessment was found to be not human subjects research. Further, all country-level data used were with the express permission of the Ministries of Health of countries involved. All interview participants were recruited between June 2020 and August 2021. Participants were interviewed purely in their professional capacities and with verbal consent. Verbal consent was obtained for both the interview and recording prior to the start of the interview, with consent being verbally confirmed after the start of recording.

### Document review

Key Act | East program documents concerning program operations during and after the global suspension in NTD operations enacted in April 2020 were identified and reviewed. Documents included Program Semi Annual Reports, available through USAID’s Development Experience Clearinghouse [16], COVID-19 Practical Approaches documents (the development of which were a key part of the COVID-19 restart strategy and which are available through the NTD toolbox) [17], as well as internal documents such as after-action review meeting minutes, presentation slides from internal webinars on COVID-19 best practices and country specific experiences, notes from Nigeria’s community listening meetings, and minutes of Act | East chief of party meetings. Descriptions of each of these documents are provided below in the section on how Act| East adapted to the pandemic.

### Program Staff Interviews

A total of 3 focus group discussions (FGDs) with the three headquarters-based country support teams and 9 key informant interviews with chiefs of party and/or their designates (referred to hereafter as qualitative interviews) were conducted with Act| East staff. Interview and FGD guides were developed inductively by a trained researcher (ES), following a review of Act | East documents described above. The following outcome thematic areas or domains were identified: communication, partnerships, relationships, learning and adapting strategies, and the feasibility and acceptability of restarting field activities, especially from the community perspective.

FGDs were held with each of the three headquarters-based teams that support all Act | East supported countries. These teams work on program management, operations, finance, monitoring, and evaluation and collaborate daily with the country programs they support. FGDs were held in August and September of 2021 with a program manager from each team giving periodic updates to follow-up questions over the rest of the study period as needed.

In addition to interviews with headquarters-based staff, one-hour long interviews were conducted with nine staff members based in six countries (Indonesia, Philippines, Nepal, Nigeria, Mozambique, and Uganda) whose role is to support national NTD programs in program implementation. Counties were selected based on availability of staff at the time. All Chiefs of Party in these countries were interviewed with three other staff members they recommended. Interview guides were shared before the interview so that respondents could invite additional participants or prepare any supporting documentation they wished to share. All interviews were conducted via Zoom and were recorded [18]. Detailed notes were taken during the interviews with additional detail provided upon review of the recording as needed. Follow up questions and updates were periodically requested of the interviewees by the study authors through direct email communication and appended to the original interview notes.

In addition to the staff interviews described above, additional focus group discussions were held with operations and finance staff concerning COVID-19’s effects on budgeting and expenditures. After the quantitative budget data were analyzed (see below for a more complete description of budget and expenditure data available), data were presented, one country at a time, to staff supporting that country in a FGD. Each FGD included 6-9 Act | East staff, including the Chiefs of Party (COPs) and other staff from the supported country and Act | East headquarters, providing financial or program management, monitoring, and evaluation.

The purpose of the FGDs was to gather context for the qualitative data and understand differences between actual expenditures and expected costs from the mid-2020 budgeting process. We also asked for explanations of specific line-item costs, for example types of PPE, that might not appear in the standardized reporting categories in financial records. In each session, one study team member facilitated while the other took detailed notes. Sessions were conducted via Zoom and recorded. Findings were summarized into slides, which the meeting participants reviewed and approved.

### Program data

We used, with the Ministries’ permission, data routinely reported by Health Ministries to the Act | East program on supported activities including the number of activities implemented by district and whether districts obtained sufficient treatment coverage. Sufficient coverage is based on reaching disease specific targets set by WHO that range from 65 to 80%. Data on persons treated is collected by drug distributors using registers or tally sheets and survey results are collected by Health Ministry survey teams using a mix of paper and electronic data collection forms. Results are then reported through the different levels of the health system. Data on MDAs for lymphatic filariasis (LF), trachoma (TR), onchocerciasis (OV), soil transmitted helminths (STH) and schistosomiasis (SCH) was included as well as data on LF and trachoma impact assessments, for the time period of October 2018 to September 2021, from eight countries.

When districts were identified as not having implemented planned activities, Act | East team members who had in-depth knowledge of country level activities were asked to provide reasons for cancellations or postponements.

### Budgets and Expenses

We analyzed Act | East country activity budgets for the program’s fiscal year (FY) 2021 period, which ran from October 1, 2020 to September 30, 2021. The analysis began in early 2021, which was after field activities had restarted but before finalized expenditure records became available. FY21 activity budgets were compared to the corresponding activity budget of a recent “typical” year, which was defined as the last year that Act | East had financial data for the same activity type (e.g. mass drug administration or survey). Data from eight Act | East-supported countries were included: Ethiopia, Indonesia, Mozambique, Nepal, Nigeria, Philippines, Tanzania, and Uganda. Data was excluded from the Democratic Republic of the Congo due to Act | East close out and four other countries for which there was not sufficient pre-pandemic comparison data (Haiti, Bangladesh, Laos, and Viet Nam). Act | East budgets were organized across countries using the same template and the same expense categories. Expenditure data were collected and analyzed once final expenditures for the fiscal year were available.

### Data management and Analysis

Data collected through document review and qualitative interviews were compiled and imported into Atlas T.I. v. 8. A thematic analysis was done using both inductive and deductive coding within pre-identified outcome thematic areas [19]. Qualitatively reported outcomes were verified using at least 2 pieces of source material.

Budget data were imported into a master Excel form and tagged according to activity and category. Simple descriptive statistics were used to compare across years. Statistical tests were not used as the quantitative review’s focus was to provide numbers for the FGDs to respond to, rather than measure any statistically significant findings. Once expenditure data were available, all results based on an analysis of budget data were verified using actual expense data in Excel.

Program data on activities were collected using Excel workbooks while routine data cleaning was managed using SAS version 9.4 [20] Analyses, such as descriptive statistics including sum, mean, and range, were also conducted using SAS. Because the data collected is reported for the universe of persons treated, and not based on a sample, statistical tests are not applied.

## Results

### How did Act | East adapt to the COVID-19 pandemic?

#### What processes were put in place to support decisions on when to restart activities?

Act | East worked closely with Health Ministries and USAID to restart Act | East support for the activities that had been suspended. The Act | East also provided support when needed on planning for restart. This process was facilitated by the development of a restart package template used to qualitatively collect and document relevant factors including: the COVID-19 conditions, national level responses, status of national government decisions on allowing restart, and descriptions of proposed modifications to reduce risk. These documents were also used to share information with USAID mission teams and RTI leadership. As time went on, and everyone became more familiar with the new approaches needed, the process was simplified. By March 2021, all Act | East-supported countries had a documented approach outlining how to conduct MDA and survey activities safely (See Table 1)

#### What adaptations were made to mitigate the impact of COVID-19 at the community level?

Safely restarting MDAs and survey activities focused on putting in place measures promoting physical distance, hygiene, and face coverings. This necessitated many changes to the delivery, training, and social mobilization strategies normally used. The change process was led by the Health Ministries in each country and the policies and standard operating procedures (SoPs) that they put in place adapted to local context. Ministries followed WHO guidance [8] and often drew on suggestions made in the Practical Approaches documents [21–23]. These Practical Approaches documents were developed when activities were suspended by national NTD programs in response to WHO guidance. Detailed descriptions were provided on how WHO guidance for implementing Neglected Tropical Diseases (NTD) Programs in the Context of COVID-19 could be applied. One document was developed for mass drug administrations, another to inform conducting lymphatic filariasis surveys, and a third on trachoma surveys.

These practical approaches were developed by a task team of NTD experts from across Act | East supported countries, which was largely seen as a benefit.

> “Act East put together a committee to put together {Practical Approaches| for our project. These were comprehensive because of drawing from so many different countries.”

#### Act | East Chief of Party

Changes made to national NTD programs by Ministries of Health ranged from small tweaks to significant redesign, necessitating large scale logistical modifications. For example, some programs changed their whole delivery strategy from a school-based model to a house-to-house model due to school closures.

Distribution of medications in communities during a pandemic is a carefully choreographed act and adaptations touched on many different elements. For example, many households were asked to provide water and soap for the drug distributors to wash their hands on arrival. Also, drug distributors found ways to reduce proximity between drug distributors and participants. They put dose poles (used to prescribe treatment dosage by height) against a wall or a tree and read it from a distance, or by they put the tablets in a bowl and then stepped back.

Training was also impacted. When training sessions could not be moved outside, larger rooms had to be found or groups split into smaller groups, requiring multiple training sessions or more trainers. Training material and social mobilization messages were adapted to cover the basics of COVID-19 transmission, review modifications, and how to communicate these to the community. Program staff reported that MDAs and training, at least initially, took longer.

Many programs reduced or eliminated the number of ‘outsiders to the community’ present for activities to reduce potential for transmission across geographies, which in turn meant a greater reliance on virtual supervision. For example, in Tanzania health officials supervised the distribution of schistosomiasis treatment by teachers remotely using photos, texts, and calls with zonal coordinators. Several programs reported that this type of supervision, which often included a wider range of players than previously, facilitated more course correction during MDA, rather than relying only on post-MDA debriefs to strategize for future rounds of MDA.

Programs also discovered unforeseen challenges to implementing modifications. Physical distancing was one of the most challenging COVID-19 protocols to implement during mass treatment campaigns and surveys as reported by Uganda, Indonesia, and Nepal teams. While physical distancing and mask use might be well adhered to during execution of program activities, this often broke down during smoking breaks, lunch breaks, and group photos. In some more remote areas, it was hard to keep the field team spaced during sleeping due to limited accommodation. Fixed posts (where the community is asked to come to specified locations at certain times) used for MDA or surveys were found to sometimes attract crowds, especially curious children, that could be hard to control. Some programs adapted to this by moving to a house-to-house model while others assigned community members to escort people from their houses to the posts in small groups. Also, early protocols had not included what to do when a team member became infected with COVID-19 - these were later developed and added.

Efforts to implement modifications also highlighted inequities and the resource-poor settings within which NTD programs may operate. For example, there was an extreme lack of safe water and or soap in some communities. Availability of face masks also varied significantly between locations. With limited additional resources, teams reported the importance of flexible budget lines to purchase necessary materials or nimbly make changes to spending plans.

#### How were adaptations developed and disseminated to facilitate global learning?

Learning what modifications were needed and whether they were feasible was an iterative process that evolved as more was known on the science of COVID transmission and as lessons were learned and shared. To facilitate this learning at program level and externally, Act| East developed a number of processes and products (see Table 2). Internally, US and endemic country-based teams debriefed initially in after action meetings following activity implementation. Key lessons learned were identified and then compiled and shared in a lessons learned slide deck and posted on Act | East’s website. Moreover, Act | East promoted direct peer–peer learning with the program. Rapid meetings were set up between country programs when lessons learned by one program could benefit another and a monthly COVID-19 Best Practice series was hosted to create a safe space to share and discuss challenges and to innovate. In addition, Act | East’s Chiefs of Party (COPs) met regularly at an RTI COP COVID-19 meeting organized by RTI’s Global Health Division, sharing their experiences and providing mutual support.

> Cross-program learning was valuable to staff, particularly early on in the epidemic, with one Chief of Party noting “The pause and reflects and COVID best practices series, especially at the beginning, were very helpful for countries and HQ staff. Those generated questions and follow ups between countries and on the chief of party what’s app group. “

As implied in this quotation, important learning also happened at country level. For example, Nigeria’s team took an innovative approach to assess community concerns using community listening sessions, implemented alongside MDA social mobilization and community education activities. These listening sessions were designed to hear and answer any concerns or questions that communities had, in an effort to increase the likelihood that communities would be willing to participate in upcoming MDA. Additionally, teams recorded community feedback as part of routine supervision during MDAs. This allowed for real time adaptation of protocols to assuage community concerns.

Other countries debriefed field staff during virtual supervision check-ins by phone or web conference to document community feedback. For example, in Haiti communities expressed increased comfort with participating in MDA because of the PPE being used. They perceived this to be more “professional” and thus more trustworthy. Another example of use of community feedback came from Mozambique where communities (in contrast to many other countries) reported discomfort with medication being dispensed from a spoon. This kind of communication between communities and the NTD program enabled the adapting of protocols to local context in real-time, improving acceptability and feasibility.

#### How did communication on the team and with other stakeholders adapt?

With movement across and within country borders heavily curtailed and in-person meetings restricted, Act | East found novel approaches to program communication both within the program and with national and sub-national partners. There was a switch to much more communication by phone, email, messaging apps like “What’s App”[24], as well as video conferencing platforms in lieu of frequent face-to-face interactions that sometimes-required long travel. This virtual communication was often supported by more thorough documentation, including minutes, transcripts, drafts, and chat logs. For some this switch in modes of communication took some time, as noted by one staff member,

> “Restrictions/lockdown means all is done by email and calls which sometimes does not carry the full richness of in person contact. Everyone had to leverage IT and there was a learning curve.”

In some cases, communications were expanded to include new people and partners. For example, in many countries there was a need to include COVID-19 committees at the district level. The strong involvement of the district health office also made the community feel more confident that the survey was safe. In other cases, communications with some partners became more frequent than before, for example, one staff member noted,

> “, [We now have] earlier and more contact with Provinces for MDA preparation. During Surveys/MDA we give time to recorders/graders to spend some time talking about COVID in the field to reassure and educate…Districts and Provinces also have ongoing COVID messages, so our practices are reinforcing MOH [COVID education]”

#### How did adaptations impact the Act East Program?

##### How did changes in communication and management approach impact program day to day operations?

One major theme emerged related to management and communication: top-down communication, management, cross country learning, and centrally coordinated policies and approaches were found to be useful at the beginning of the pandemic when so much was new and unknown to everyone. As time went on, and programs gained experience, adapting to local situations became increasingly important and top-down approaches could become cumbersome. The flexibility to adapt quickly and flexibility became increasingly important as the epidemic wore on and programs restarted activities safely and successfully.

Sharing lessons across countries from teams who were “firsts’’ was widely cited as an example of something that was helpful in the early days. For example, DRC were the first Act | East-supported country to implement a trachoma survey using innovative face shields and the Indonesia team had a strong response to the first COVID-19 case reported among team members during a community visit. Act | East’s country-led development and dissemination of the Practical Approaches documents [21–23] was also appreciated by many countries who used these to inform the development of their own documents and guidance. Similarly, the re-start packages, described above, were seen as important in providing a way forward in the early days and facilitated a rapid and safe resumption of activities. As one chief of party noted

> “, Once SOPs were safely implemented it made people more comfortable about going to the field.”

However, as time went on, both team members based in the central and country offices reported that continued sharing seemed to be of more interest and value to the central office-based team members while countries had moved more focus to country-specific operations management for restarted implementation. Similarly, the re-start packages process started to feel more burdensome once implementation restart regained momentum.

Continuing to relay messages on multiple moving pieces between program country-based teams (in regular communication with Health Ministries) and Act | East headquarters level (in regular communication with USAID), through multiple people, made for increased chances of miss-communication, longer response times, and less flexibility. Thus, in later phases of the pandemic Act | East reduced the frequency of program-wide COVID-19 specific round tables on lessons learned, countries relied on their own mitigation measures and documents, and the restart process adapted first to a much simpler restart package option and then to short monthly updates. One Act | East staff member reported,

> “Approval and review [for restart] was clunky but has definitely become more streamlined [since restart packages were adapted] …… [It was] much more burdensome in the beginning.”

Greater flexibility was operationalized in concrete ways. For example, budgeting for meetings allowed enough resources for either virtual or in-person meetings, trainings, and workshops. More resources were also freed up when certain supplies (such as PPE) became easier and cheaper to procure. The ability to pivot and reprogram resources quickly under a certain threshold allowed for a last-minute pivot should local COVID-19 conditions changes. Local adaptations to implementation procedures were also made, such as a lasting shift to more door-to-door MDA, a greater emphasis on pre-MDA planning with district officials. While this additional pre-MDA planning was necessary to ensure safety during the height of the COVID-pandemic, other benefits, such as wider use of micro-planning and pre-MDA censuses have accrued from this additional planning effort.

Throughout the restart of community-based NTD services and data collection, and as the number and frequency of community-based activities continued to approach pre-pandemic levels, program staff at the partner, country, and subnational level noted that there were many tradeoffs related to changing communication channels, some of which are summarized in figure 1 These tradeoffs represent a recognition that adaptations to pre-pandemic communication patterns came with both pros and cons from the perspective of effective NTD program administration and service delivery.

**Figure 1.**
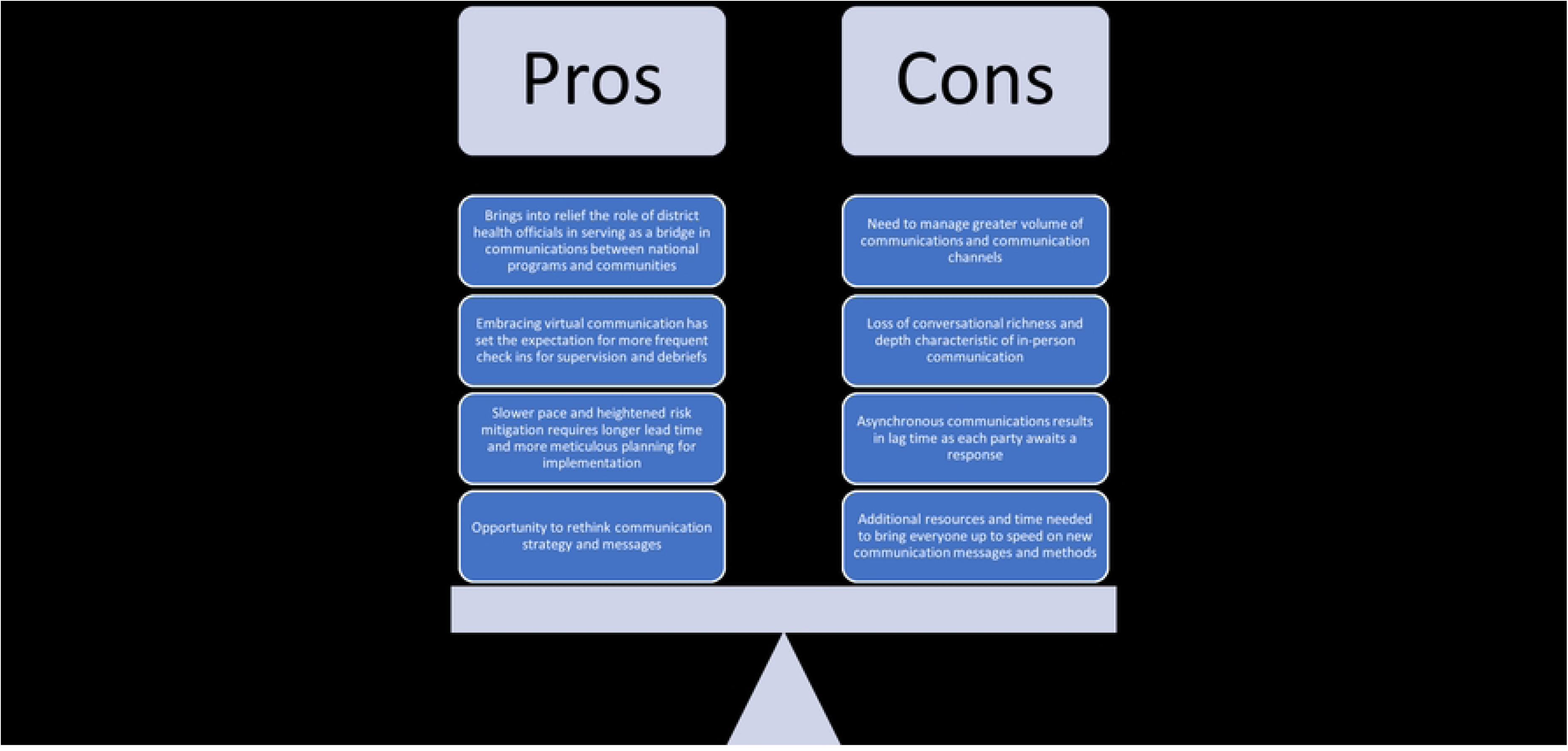
Tradeoffs associated with changing communication norms during the COVID-19 restart process.

##### How was the quality of partnerships impacted?

Another strong theme was the impact responding to the pandemic had on program partnerships. For Act | East, key relationships include those with the Health Ministries, with the donor (USAID), among consortium organizations, and among program functional teams.

Having strong relationships with Health Ministries going into the pandemic and clear ongoing communication during the pandemic was reported as key. This helped weather the short-term strain put on relationships for example when one organization resumed in person working before another, or when differences in mitigation policies (e.g. around maintaining physical distancing and wearing face coverings) started to emerge.

> “Strengthening partnerships and having functional relationships with the Ministry is very helpful [during restart planning].” Act | East Chief of Party

Act| East endemic country teams reported more engagement with both program headquarters-based program staff and with USAID. On the one hand, this offered them greater insight and participation in decision making and new opportunities for leadership roles. However, this increased engagement, along with increased expectations for documentation, added to the workload and stress. USAID country missions also became more involved during the restart process, coordinating information and responses across USAID-funded programs.

#### How did COVID impact NTD program coverage?

##### How were program modifications perceived by communities?

A critical component of community-based service delivery programs is the trust communities have in the safety and value of the services [25]. The success of resuming MDAs and disease surveillance activities during the pandemic was contingent upon the acceptance from the community. Two of the earliest countries to resume community-based activities were the Democratic Republic of the Congo (DRC) and Mozambique. These countries included brief household surveys as part of house-to-house trachoma surveillance surveys [12]. These surveys revealed a high level of comfort of households with survey teams (a comfort level of 99-100% in both countries, according to supervision checklist data) in their compounds and high level of assurance with the COVID-specific adaptations that were made by survey teams. This high level of comfort was reassuring to the NTD community, especially as trachoma disease surveys include an eye exam requiring close physical contact with household members. However, experiences were varied. One Act |East staff member noted that,

> “Communities were ready to collect what they called ‘eye medicine’…they trust their community health workers and what they have to say about MDA [safety]”

 while another staff member pointed out that

> “It is taking more time than expected to get local communities on board for activities… flexibility is key and it is important to work with districts and communities to get them on board.”

### What percent of the MDAs and surveys planned between October 2018 and September 2021 were implemented and why were activities not implemented?

As noted above, the WHO recommendation pausing all NTD MDA and survey activities issued on April 1st, was lifted on July 27, 2021. They recommended that countries restart activities only after careful consideration of the relative risks of restarting versus remaining in suspension and putting in place mitigation measures that would reduce likelihood of transmitting COVID-19. This suspension in activities obviously impacted the ability of NTD programs to implement planned activities. Total numbers of MDAs and DSAs planned and implemented by disease are presented along with main reasons given for cancellation or postponement of planned activities as the overall patterns were similar for all.

COVID-19, which hit in the middle of the program’s 2020 fiscal year (FY) had a significant impact that year (see figures 2 and 3). Out of 748 disease-specific MDA planned in 10 countries, only 276 (37%) were implemented, compared with more than 90% in both FY18 and FY19. 59% (154) of these were implemented before COVID-19 was declared a global pandemic. COVID-19 was the reason given for not implementing MDA that year in 86% of cases. it is estimated (based on the expected numbers of individuals targeted for treatment during that time frame) that 38,928,375 individuals were affected by COVID=19-related MDA cancellations.

**Figure 2.**
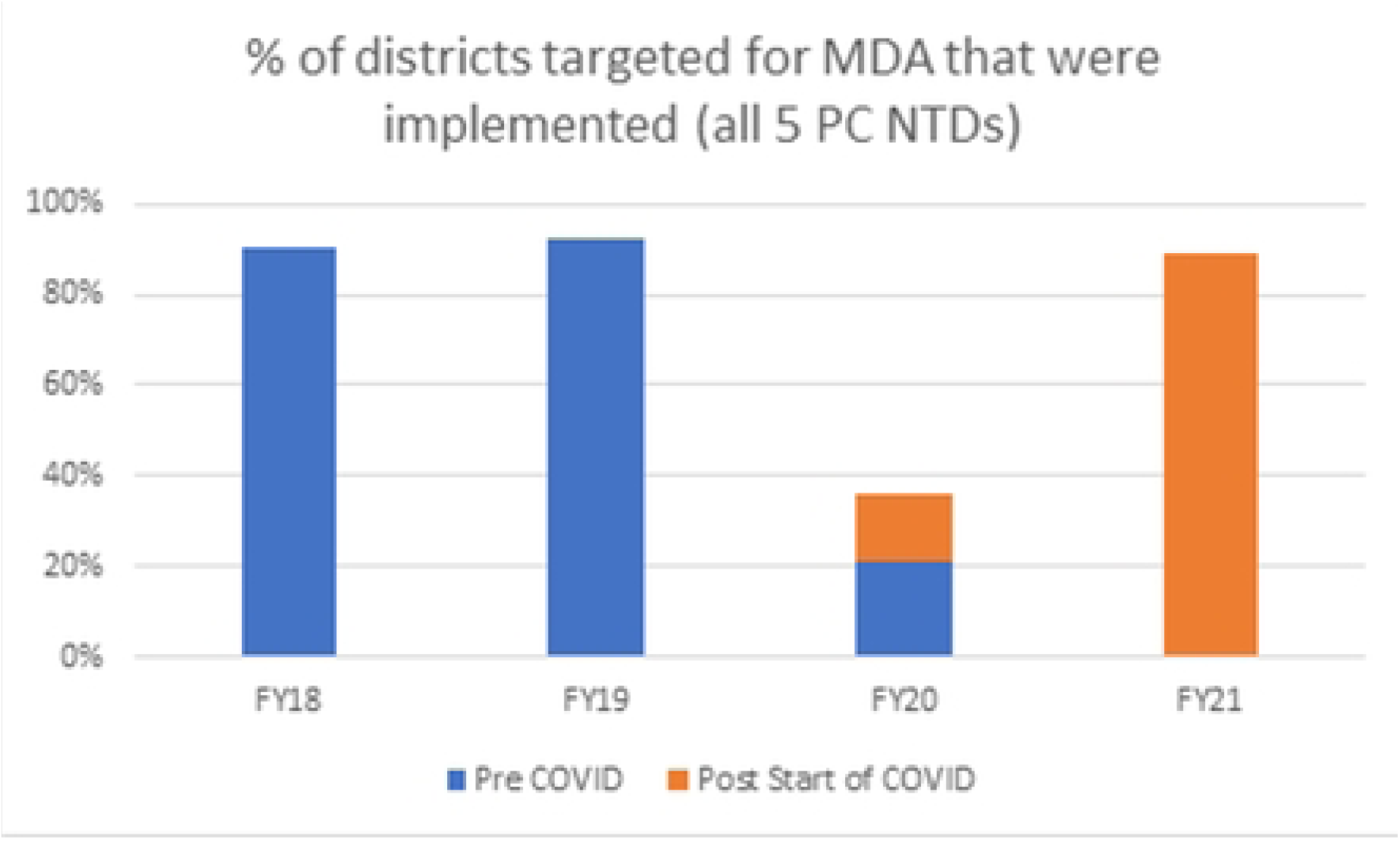
Percentage of all districts that were anticipating MDA (for any of the 5 preventative chemotherapy neglected tropical diseases) where MDA was successfully implemented, by fiscal year

**Figure 3.**
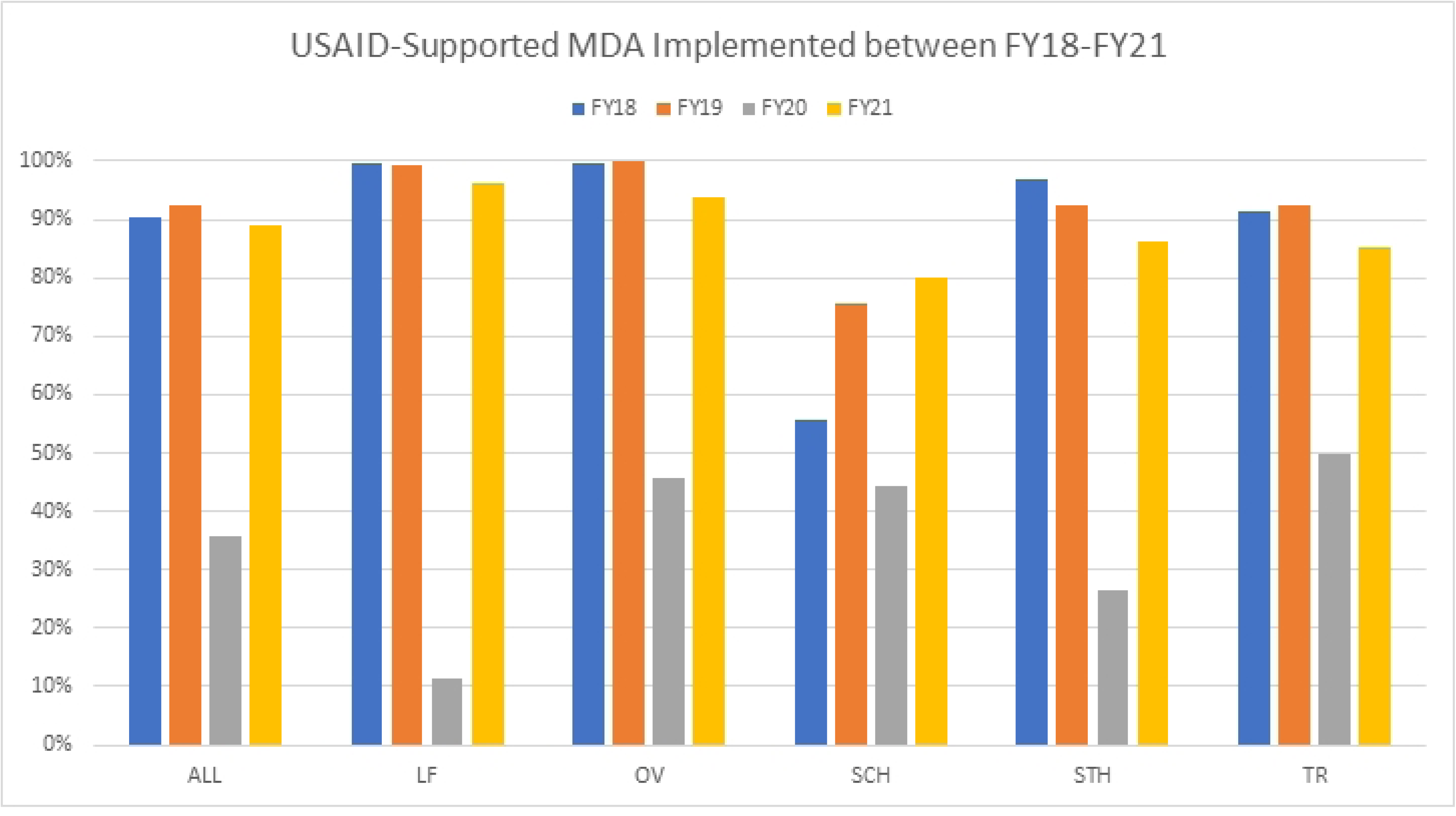
Percentage of districts anticipating MDA, that received MDA, by disease and fiscal year

This turned around quickly again in FY21, with 88.6% (669 of 755) of planned MDAs implemented (figures 2 and 3). Of the 86 district MDAs not implemented, COVID-related delays were no longer cited for lack of implementation. The most important issues featured, instead, were unrest/insecurity, lack of drugs unrelated to the pandemic and pending survey results to confirm whether MDA was required.

As Ministry of Health programs get closer to elimination of certain NTDs, more districts move into the surveillance phase, and the number of planned surveys increases. The impact of COVID-19 on these has been significantly greater than MDA (see Figure 4). In FY21 only 41% of LF and Trachoma surveys planned were implemented compared to 74% and 73% in the 2 years pre-COVID.

**Figure 4.**
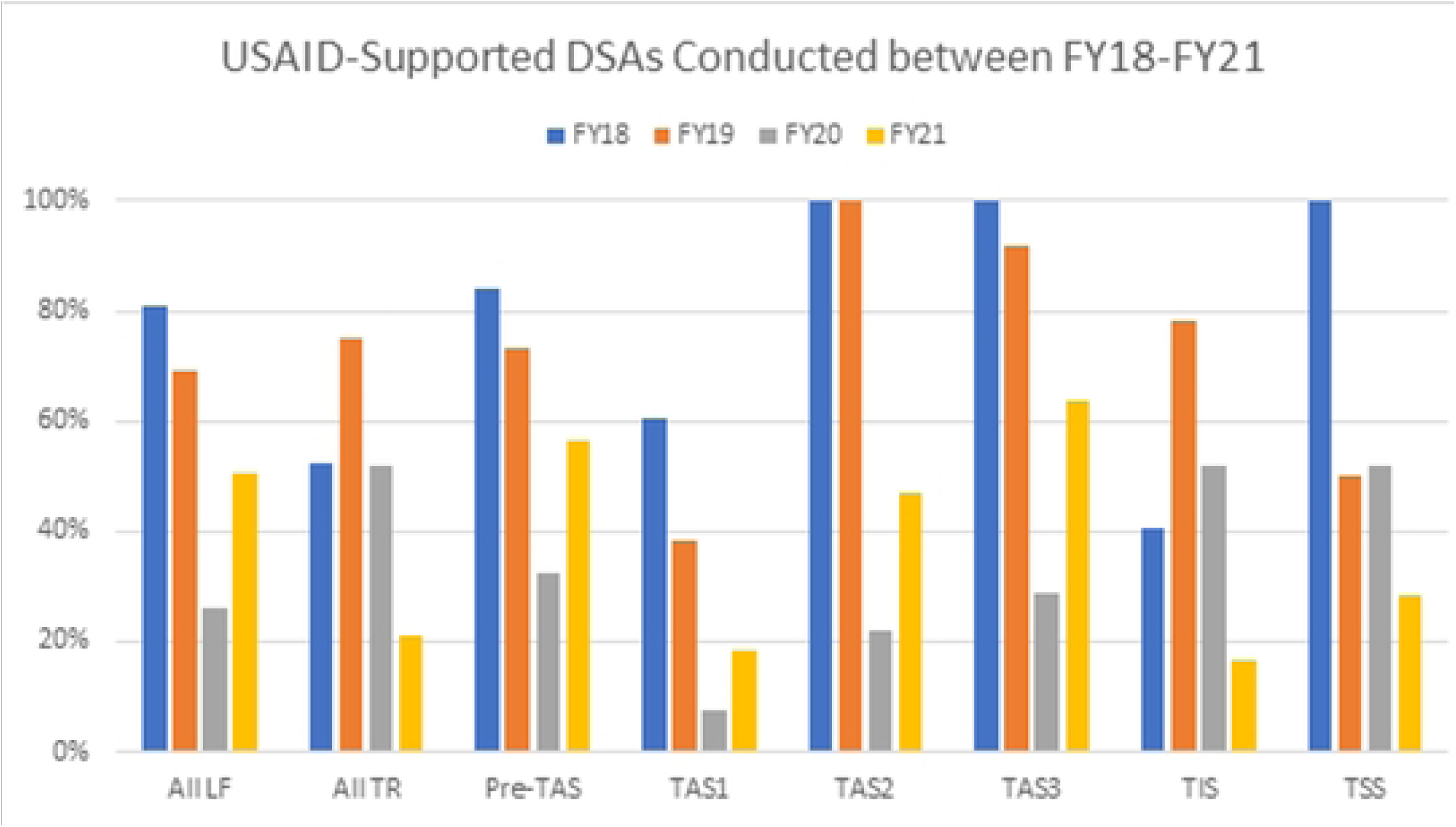
Percentage of districts planning a Disease Specific Assessment (DSA), that implemented a DSA, by disease and fiscal year.

The difference in restart between MDA and survey activities reflects at least in part the fact that MDAs were prioritized for restart over surveys due to the impact on NTDs of delayed MDAs being greater. Additionally, the program took extra precautions before restarting survey activities to mitigate the potential increased risk of survey teams becoming infected with COVID-19 because of the physical proximity experienced during blood draws and eye examinations.

### What was the change in treatment coverage?

Throughout this period, treatment coverage, measured as the percent of districts reaching sufficient coverage, also remained high. In the two years pre COVID, 93% and 92% of district level MDAs supported, across 13 countries, achieved sufficient coverage. In FY21, 82% achieved sufficient coverage (see Figure 5).

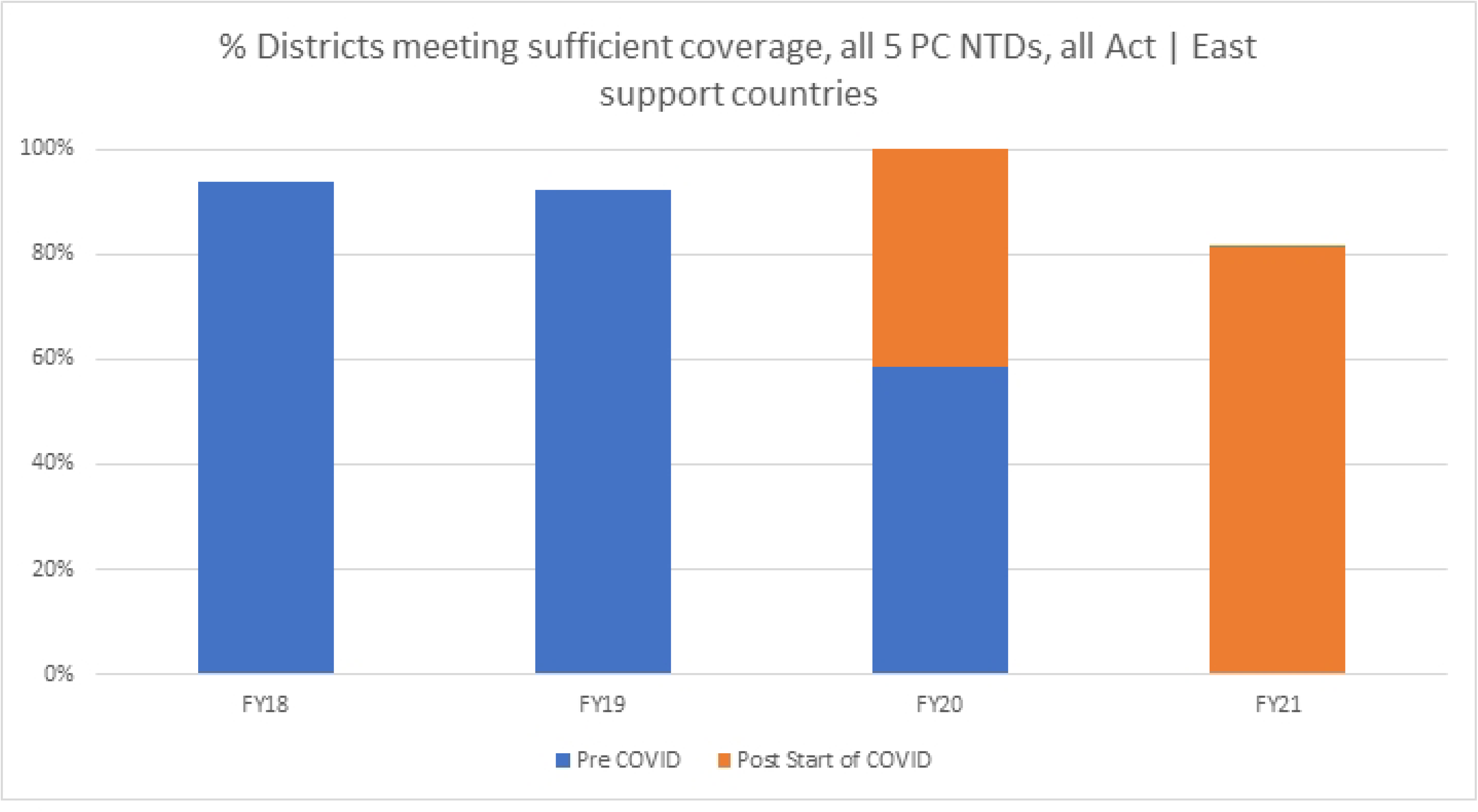

### How did COVID impact program resources?

#### How did COVID-19 affect the cost of implementation?

The effects of COVID-19 on resource use varied widely by country. The change in budgeted MDA costs per district varied from a 71% decrease to a 20% increase; budgeted DSA costs per survey decreased by as much as 87% in some countries and increased by as much as 119% in others. The analysis of expenditure data revealed similarly wide rages of results. The extreme variation in changes of costs by country led the study team to conclude that a single global figure could not convey the results of COVID-19. Instead, the value of understanding cost implications came from understanding individual country contexts. We therefore used the quantitative figures to guide qualitative data discussions; we draw the results presented here primarily from focus group discussions.

In terms of resources outside of the program’s financial records, focus group participants expressed concern about the intangible costs to NTD programs’ momentum, as well as the weakening of interpersonal relationships due to the shift to virtual meetings. Many teams also reported that the national government had shifted funds and staff members from NTD work to the COVID response. However, our analysis of budgets and expenditures have shown that the reallocation and reprioritization of national NTD program budgets has continued three years after the onset of COVID-19, with varying success in advocacy for integration or reverting to original NTD program budgets.

#### What drove changes to costs?

Despite the broad ranges in percent change to budgeted costs, and the importance of contextualizing budget figures, however, some key messages were repeated across countries through FGDs. In some countries, the variability in costs derived largely from changes in delivery platform, such as moving from school-based to community-based distribution due to COVID school closures. Travel costs faced two opposing forces: a decrease in costs due to travel restrictions and/or improved up-front logistics planning versus an increase in costs due to social distancing requirements. These requirements mandated the rental of additional vehicles and hotel rooms, often at least doubling the associated costs. Fuel costs also increased more rapidly than expected during the pandemic. Supply costs generally increased, with infection prevention materials added to budgets. In addition, the predictability of supply costs waned due to variable inflation rates and fluctuating product availability. As one qualitative interviewee noted “When we assembled the budgets, we didn’t know exactly what would be required. For example, would we need to give masks to everyone? It turns out that they only needed to give masks to the staff and volunteers, not the general public.”

The financial costs of meetings decreased due to the shift to virtual meetings. Some country teams expressed concern that this financial savings came with a loss of meeting effectiveness, though others praised the increased accessibility of virtual meetings. In general, focus group members thought that the financial costs of COVID would recur in future years of the pandemic. They emphasized, however, that the variability and uncertainty of the required costs would likewise continue. This is illustrated by the staff member who noted,

> “There will be contingencies or immediate needs that arise not in the [pre-covid] workplan… so there is a need to have flexibility in workplans and budgets to respond to these contingencies.”

## Discussion

This study provides a critical analysis of a large and complex global health program’s experience of managing amidst the uncertainty caused by COVID-19. The program studied was designed to support the elimination and control of five different NTDs in thirteen countries. COVID-19 resulted in most of the supported activities, including mass drug administration and disease assessment surveys, being stopped for four months, followed by a period of slow restarting with adaptations made to reduce the risk of COVID-19 transmission. While other published articles have described adjustments and innovations made to get NTD programs up and running again, this paper is unique in its use of outcome-harvest methodology, a complexity-aware technique, that enables an analytic exploration that goes beyond the rich description on what was done to assess the economic and financial costs, challenges, and successes associated with adapting program operations. It does so by considering the experiences of ten countries and goes beyond the initial emergency response period to the day-to-day of living with COVID-19 and triangulates programmatic data and costing reviews with qualitative interviews that introduce a diversity of perspectives.

This study identified several processes and tools developed by the program to manage the initial challenges brought on by the pandemic. Two of these processes were the ‘restart package templates’ and the “Practical Approaches documents”. Both of which were developed while community-based service delivery was suspended. Other papers published have described similar approaches to the ‘restart package templates’ designed to facilitate discussions on when and how to restart programs during the COVID-19 pandemic including the Risk Assessment and Mitigation Action (RAMA) tool and a matrix tool [9–14] The templates described here, while covering similar issues, differed in that instead of providing a score on each issue, they focused on collecting qualitative information to support decisions on restart readiness. The qualitative element was especially useful at the beginning of the restart process when information was fragmented and communication between stakeholders was critical.

Other adaptations Act | East found valuable included concrete tools and guidance such as modified supervision checklists, training materials, and clear guidance on policies and procedures around convening, traveling, testing, and so on. While each country’s government was responsible for the development of national SoPs, the global guidance provided by the WHO and the information in the Practical Approaches documents supported planning.

Additionally, while program communications moved to more frequent, problem-focused virtual communication, the ability to share innovations and experiences with restart raised morale and confidence during the restart process. One Act | East modification of facial shields that could be used for trachoma eye exams which raised the comfort of programs returning to trachoma activities [12]. These findings add to the growing literature which has also emphasized the importance of supervision checklists, PPE trackers, clear standard operating procedures, and frequent virtual communication and coordination at all levels [9–12]. That being said, however, our experience in implementing across country contexts was that these tools and guidance were only as useful as the ability to be flexible in their application. The situations changed so frequently (what venues were open, what the government would allow, next steps if a staff person tested positive for COVID-19 during implementation) that structured tools coupled with flexibility in special instances were really the difference maker in allowing for a successful restart.

The restart, however, was not without its challenges. In line with previously published studies, we encountered challenges early in the procurement of PPE (Personal Protective Equipment), PPE compliance, access to handwashing facilities, adherence to proper handwashing practices, and increased time needed for training and implementation [9–11,14]. Others have cited misinformation about COVID-19 amongst the community as a barrier to entering and/or interacting with those they were providing with NTD treatment.^26^ However, Act | East implementers did not face a significant amount of COVID-19-related community resistance. This could be attributed to a long history of implementation in endemic communities in these countries and further attempts to build community trust through COVID-19-sensitive social mobilization and community listening sessions. Where applicable, Act | East’s strong partnerships, with government, community structures, or other implementing organizations also contributed to limited resistance. This finding emphasizes the importance of having established community trust and good-will in early responses to a public health emergency.

Another area of adjustments made during the early restart process included planning and budgeting for COVID-affected activities. Practical changes to approved workplans and budgets had to be made to account for new expenses such as PPE, upgraded technology and internet to support virtual communication needs, and a move to house to house mass drug administration to reduce potential crowding at fixed-post-delivery sites. Act | East finance and implementation teams also noted several changes to field preparation that were likely to continue if the COVID-19 emergency was ongoing. Some of these future considerations, such as hiring more vehicles to account for physical distancing, capacitating health workers to educate communities on COVID-19, holding trainings outdoors or creating ventilation indoors, reducing the training class size per event, increasing the number of days to implement during COVID, and ensuring access to water or sanitizer to promote good hygiene practices amongst implementing teams, have also been noted by other NTD implementation programs [10,11,27].

Over time, as more NTD control and elimination efforts were restarted, the uncertainty on whether and how to implement safe and effective NTD program activities was superseded by questions regarding how programs could better manage the complexity of managing a “new normal” in which COVID-19 will continue to affect countries in waves, requiring consistent learning and adapting to continue to progress towards NTD elimination and control.

As predicted by organizational and management theory as applied to disaster management, while a “task-oriented” hierarchical management structure is fundamental to an initial response to an emergency, a shift to a more decentralized and matrixed organizational and management structure is needed as the initial response moves into recovery and mitigation phases [28]. Similarly, the initial creation of practice recommendations and approval of structured restart packages in the initial stages of the COVID-19 pandemic gave way to more flexible, adaptive management approaches at the country level as the pandemic wore on. For example, the activity approval processes were abbreviated and streamlined after the first 6 months of successful implementation of community-based service delivery after restart. Local adaptation of SoPs and procedures have similarly been adjusted as COVID-19 testing and vaccines have become available or as additional waves of infection have prompted localized shutdowns.

An important lesson learned is the importance of nimble programming that can continuously adapt to change within a relatively short term. The flexibility to make rapid pivots and continuously re-program and re-allocate resources helps mitigate uncertainty by preventing programmatic paralysis. Activities can be adjusted in near-real time as local conditions allow. However, this flexibility can also come at a cost. SoPs enable consistent quality of service delivery. Also, last-minute activity shifts can impact efficiency through additional resource requirements needed to accommodate a shift in planned activities. The former concern can be partially ameliorated through intentional attention to program monitoring and supervision. It is also especially critical that collaboration, learning, and adapting (CLA) approaches are incorporated into program monitoring and supervision structures since conditions can change rapidly and pragmatic adaptations and solutions can become part of the pool of resources available to the program world-wide. This attention to careful process monitoring and collaborative learning and adapting can allow for evidence-driven program adaptation, even over the short term.

An important additional lesson learned from the Act | East experience is the understanding that, in many cases, contingencies considered during the operational planning and budgeting phase can offset the latter concerns around resource requirements. While programs want to have a detailed budget and activity plan for good stewardship, systems need to have enough flexibility to adapt to changing circumstances. In Act | East, this was achieved in part through setting thresholds for who can approve certain types of changes to budgets/activities so that small budget changes can happen more quickly while changes to deliverables were agreed upon on a country-by-country basis. This flexibility helped to overcome budgeting concerns associated with supply chain issues, temporary lockdowns, travel restrictions, and other disruptions to budgets and implementation associated with COVID-19 that were recognized very early in the pandemic [10,33].

Overall, program restart during the pandemic generated many changes to how the program supports country NTD programs and added new operational and relational norms and processes for donors, partners, and governments. Still, fears at the start that program services would stall, diseases resurge, and that the increased workload that accompanied program restart might continue indefinitely have not been borne out in Act | East supported countries and communities [4–7]. Once restarted, mass drug administration and disease surveillance in the 8 countries we examined were resumed with a pace and quality consistent with pre-COVID levels.

NTD modelers have since estimated that the potential for NTD resurgence in response to disruptions is slow compared to malaria, measles, and other infectious diseases. Trachoma, schistosomiasis, and soil transmitted helminthiasis face a greater risk of resurgence compared to lymphatic filariasis and onchocerciasis [28–29]. In all five of the diseases, the longer the interruption without intervention, the greater chance and higher rate of resurgence and resurgence is likely to happen more rapidly in high transmission areas [29–30]. To respond to these predicted setbacks, the modeling community highlighted some suggestions to maintain high quality programming and to “catch-up.” Where possible, they recommended finding ways to increase coverage and / or ‘catching up’ by increasing frequency of MDA biannual MDA, increased mitigation strategies for SCH and trachoma including extending coverage and treatment to adults [28,31,32]. These recommendations do not come without additional costs of their own.

While it seems that long term disruptions to Act | East programming have not materialized, we recognize that Act | East’s experience does not necessarily speak to all NTD programs, everywhere. Act | East benefitted from working with governments that had well-established programs and having long term relationships with these. Another interesting observation is that many of the changes to operations and ways of working that have contributed to this successful restart have become normalized. For example, many programs have continued to include more house to house, rather than fixed-post distribution models for mass drug administration.

This success provides important lessons for ongoing program support during future emergencies that carry the potential to disrupt community-based service delivery for NTDs or other essential primary health programming. Having some structured guidance that is heavily leveraged by flexible and responsive systems was critical for the success of Act | East program restart during the COVID-19 pandemic but is also important from a future-proofing perspective. Uncertainty is a certainty, and it can be expected that in the next crisis, some of the specific adaptations and solutions that were helpful during program restart during COVID-19 will be useful again. Inevitably, however, some of these specific adaptations will not speak to the unique characteristics of the next emergency. Having a template for how community and epidemiological assessments can be made and how responses can be tailored to unique contextual factors will allow each new emergency to be addressed through a timely and practical approach.

With an eye towards the future, there are several adaptations to Act | East’s programming that were undertaken during COVID-19 restart that have been and expect to continue to be maintained in the post-pandemic era. Given the need discussed above for strengthened supervision and monitoring, the program has invested in increased real-time supervision tools including electronic data collection, dashboards, and virtual supervision support. There has also been a greater emphasis on closer coordination with and empowerment of subnational implementation teams to carefully plan the details of mass drug administration or household surveys well in advance to ensure all COVID-19 protocols are adhered to during activity implementation. Finally, there has been a real focus on safety and fidelity to process and protocol over concerns about scale and economy. These adaptations have collectively led to an increase in data quality across the program.

The additional need for careful coordination and the added emphasis on the value of sharing lessons learned program wide have also manifested in strengthened communication across the program and between partners. In many cases new partnerships and collaborations were formed and in others existing relationships were strengthened by the need for greater communication and collaboration both from the “top down” as well as the “bottom up”.

Act | East also found that the COVID-19 restart sped up the leveraging of information technology solutions. These solutions facilitated those improvements in monitoring and supervision as well as the strengthened communication and collaboration described above by enabling more virtual data collection, real-time remote supervision, and the use of diverse technology platforms for communication and learning. While in some cases the use of technology saved money and time for meetings, transport, and training, the greater emphasis on safety and more careful planning also came with a requirement for more resources to safely implement, particularly in terms of material costs, staff time, and planning timelines.

While the study draws on several strengths, it is not without its limitations. While the data were collected prospectively, that is concurrently with program operations, the analysis was retrospective in nature and the qualitative data, could be biased with the clarity of hindsight.

Further, given the intense scrutiny that country programs were experiencing, especially early in the pandemic, the safe implementation and high coverage rates that country activities reported could have been influenced by observation bias. It also must be stated that, while this study sought out the perspectives of multiple stakeholders, many of the adaptations and learnings are filtered through the lens of implementing partners who are supporting multiple country programs and that many of the learnings reported were learned through trial and error at the national and sub-national levels. Both the implementing partner and many country programs benefitted especially from those country NTD programs that were the earliest leaders in NTD program restart during COVID-19 as a result. Further, it is important to note that while all countries have experienced COVID-19, each country operated to varying degrees and on different timelines across the globe. There was no true comparison group since the pandemic was truly global and no country could claim to be completely unaffected by COVID-19. Finally, several of the included countries had NTD activities supported by other partners which fell outside this study’s scope. Further, no intention is made to generalize Act | East’s experience and perspective to all implementing partners, country programs, or donors.

Given early modeling that predicted that gains made by NTD programs worldwide would be eroded by a prolonged disruption in programming, strong efforts were made by NTD programs to restart mass drug administration and disease surveillance as soon as possible. This paper contributes to the growing literature on early adaptations made and provides further insight to longer term adaptations that resulted and of the impact that pandemic-inspired adaptations had on partner relationships, cost, and program coverage. Many questions remain about how the changing faces of the COVID-19 pandemic continues to impact NTDs, especially as COVID-19 intersects with other emerging emergencies such as the recent global outbreaks of Monkeypox[24], Ebola [35], Cholera [36], Polio [37], and 2022’s “tripledemic” of influenza, the Respiratory Syncytial Virus (RSV), and COVID-19 [38] that struck North America and the European Union. As many countries approach the “last mile” towards the elimination of certain NTDs, further consideration should be given to the impact that public health emergencies will have on progress towards elimination. These findings demonstrate that, even in the face of a global health security crisis, that it is still possible for resilient and adaptable health programs to maintain momentum in essential public health program areas such as neglected tropical diseases.

## Data Availability

All data presented are the owned by the ministries of health whose countries are included in this analysis. All data included in this paper are used with express permission of the data owners.

## Acknowledgements

The authors would like to acknowledge the leadership and staff of USAID, Act | East, and the country ministries of health, particularly the NTD and Infectious Disease staff whose collective efforts are what made the program adaptations and effects possible. The authors would also like to extend thanks to the communities who welcomed NTD programs back to their communities in a time of uncertainty.

---

| High Level Question | Sub Questions | Data Sources |
| --- | --- | --- |
| 1)How did Act \| East adapt to the COVID-19 pandemic? | What processes were put in place to support decisions on when to restart activities?<br>How did communication on the team adapt to lockdown?<br>What adaptations were made at the community level to mitigate the impact of COVID-19 and how were these developed, disseminated, and implemented? | Practical Approaches Documents<br>After Action meeting notes<br>Lesson learned slide deck<br>Peer-to-peer learning session notes<br>COP COVID-19 meeting minutes<br>COVID-19 Best Practice Series presentations<br>Qualitative interviews with Act\| East project staff |
| 2)How did adaptations impact the management of the Act East project? | How did COVID-19 adaptations impact program day-to-day operations?<br>How was the quality of relationships and partnerships impacted? | Practical Approaches Documents<br>After Action meeting notes<br>Lesson learned slide deck<br>Peer-peer learning session notes<br>COP COVID-19 meeting minutes<br>COVID-19 Best Practice Series presentations<br>Qualitative interviews with Act\| East project staff |
| 3)How did COVID-19 impact NTD program coverage? | How were program modifications perceived by beneficiaries?<br>What percent of the MDAs and surveys planned between October 2018 and September 2021 were implemented and why were activities not implemented?<br>What was the change in treatment coverage? | Household questionnaires included in supervision documents<br>Field and supervision notes from restarted social mobilization, mass drug administration, and disease specific assessment activities in endemic communities<br>Qualitative interviews with Act\|East project staff<br>Act\|East Semi-Annual Reports<br>Act\|East project monitoring data<br>Health Ministry data routinely reported to Act\|East |
| 4)How did COVID impact program costs? | How did the impact impact the cost of implementation?<br>What drove changes to costs? | Act\|East project budgets<br>Act\|East project expenditure data<br>Qualitative interviews with Act\|East project staff |

| Product/process | Description/Purpose |
| --- | --- |
| Lesson learned slide deck | To gather and disseminate common experiences and outcomes from initial implementation efforts in the COVID-19 environment, lessons learnt were gathered and shared in a slide deck available publicly on the Act \| East website. |
| After Action meetings | These were held internally within the project to informally debrief within a few days of each countries' first implementation of an activity in the COVID-19 context. Project country teams presented the activity and lessons learned via powerpoint to a small home office team. Key lessons were captured, disseminated across the project. |
| Country - to- Country ad hoc meetings | Rapid meetings were set up to informally share lessons learned between specific country teams facing similar situations. |
| COVID-19 Best Practice Series | More formal sharing of lessons learnt across the project took place in the "COVID Best Practices Series". All team members were invited and staff from different countries took turns presenting their experiences and lessons learnt. This was seen as a 'safe space' for individuals to ask questions and provide feedback and suggestions. |
| COP COVID-19 meeting | These were routinely held by RTI's global health leadership, during the first few months of the pandemic, to facilitate sharing experiences between countries and across projects. COPs from multiple projects met weekly to share experiences including how to lead their teams at this challenging time and on making implementation and restart decisions. Meetings became less frequent (monthly and then quarterly) as the study period progressed. |

## References

1. David Williams O, Yung KC, Grépin KA. The failure of private health services: COVID-19 induced crises in low- and middle-income country (LMIC) health systems. Global Public Health. 2021/09/02 2021;16(8-9):1320–1333. doi:10.1080/17441692.2021.1874470

2. World Health Organization. WHO issues interim guidance for implementation of NTD programmes. 2020.

3. World Health Organization. WHO Coronavirus (COVID-19) Dashboard. https://covid19.who.int/

4. Abdela SG, van Griensven J, Seife F, Enbiale W. Neglecting the effect of COVID-19 on neglected tropical diseases: the Ethiopian perspective. Transactions of the Royal Society of Tropical Medicine and Hygiene. 2020;114(10):730–732.

5. Molyneux DH, Aboe A, Isiyaku S, Bush S. COVID-19 and neglected tropical diseases in Africa: impacts, interactions, consequences. Oxford University Press; 2020. p. 367–372.

6. Ung L, Stothard JR, Phalkey R, et al. Towards global control of parasitic diseases in the Covid-19 era: One Health and the future of multisectoral global health governance. Advances in Parasitology. 2021;114:1–26.

7. Prada JM, Stolk WA, Davis EL, et al. Delays in lymphatic filariasis elimination programmes due to COVID-19, and possible mitigation strategies. Transactions of the Royal Society of Tropical Medicine and Hygiene. 2021;115(3):261–268.

8. World Health Organization. Considerations for implementing mass treatment, active case-finding and population-based surveys for neglected tropical diseases in the context of the COVID-19 pandemic: interim guidance. World Health Organization Geneva; 2020.

9. Amanyi-Enegela JA, Burn N, Dirisu O, et al. Lessons from the field: delivering trachoma mass drug administration safely in a COVID-19 context. Transactions of The Royal Society of Tropical Medicine and Hygiene. 2021;115(10):1102–1105.

10. Kabore A, Palmer SL, Mensah E, et al. Restarting neglected tropical diseases programs in West Africa during the COVID-19 pandemic: lessons learned and best practices. The American Journal of Tropical Medicine and Hygiene. 2021;105(6):1476.

11. McKay S, Shu’aibu J, Cissé A, et al. Safely resuming neglected tropical disease control activities during COVID-19: Perspectives from Nigeria and Guinea. PLoS Neglected Tropical Diseases. 2021;15(12):e0009904.

12. McPherson S, Stern J, Bush S. Innovative tools to advance trachoma elimination in the context of COVID-19. Community Eye Health. 2021;34(111):28.

13. Molyneux D, Bush S, Bannerman R, et al. Neglected tropical diseases activities in Africa in the COVID-19 era: the need for a “hybrid” approach in COVID-endemic times. Infectious diseases of poverty. 2021;10(01):74–86.

14. Sakho F, Badila CF, Dembele B, et al. Implementation of mass drug administration for neglected tropical diseases in Guinea during the COVID-19 pandemic. PLoS Neglected Tropical Diseases. 2021;15(9):e0009807.

15. Wilson-Grau R, Britt H. Outcome harvesting. Cairo: Ford Foundation. 2012;

16. USAID. Development Experience Clearinghouse. https://dec.usaid.gov/dec/home/Default.aspx.2023.

17. RTI International. Practical Approaches to Implementing WHO Guidance for Neglected Tropical Disease (NTD) Programs in the Context of COVID-19: Mass Drug Administration (MDA). 2020. RTI International. NTD Toolbox. https://www.ntdtoolbox.org/

18. Zoom Video Communications. Version 5.13.10. 2011.

19. ATLAS.ti. Version 22.0.6.0. 1993.

20. Statistical Analysis System. Version 9.4. 2020.

21. https://www.ntdtoolbox.org/COVID19MDA

22. RTI International. Practical Approaches to Implementing WHO Guidance for Neglected Tropical Disease (NTD) Programs in the Context of COVID-19: Lymphatic Filariasis (LF) Surveys. 2020. https://www.ntdtoolbox.org/COVID19LFSurveys

23. RTI International. Trachoma Surveys: Practical Approaches to Implementing WHO Guidance for Neglected Tropical Disease (NTD) Programs in the Context of COVID-19. 2020;

24. WhatsApp. Version 2.2043.21. 2022. https://whatsapp.com

25. Krentel A, Fischer PU, Weil GJ. A review of factors that influence individual compliance with mass drug administration for elimination of lymphatic filariasis. PLoS neglected tropical diseases. 2013;7(11):e2447.

26. Ahorlu CS, Okyere D, Pi-Bansa S, et al. COVID-19 related perception among some community members and frontline healthcare providers for NTD control in Ghana. BMC Infectious Diseases. 2022;22(1):1–12.

27. Amazigo UV, Leak SG, Zoure HG, et al. Community-directed distributors—The “foot soldiers” in the fight to control and eliminate neglected tropical diseases. PLoS neglected tropical diseases. 2021;15(3):e0009088.

28. Tian J, Zou Q, Cheng S, Wang K. A framework of task-oriented decision support system in disaster emergency response. Springer; 2009:333–336.

29. Hollingsworth TD, Mwinzi P, Vasconcelos A, De Vlas SJ. Evaluating the potential impact of interruptions to neglected tropical disease programmes due to COVID-19. Transactions of The Royal Society of Tropical Medicine and Hygiene. 2021;115(3):201–204.

30. Toor J, Adams ER, Aliee M, et al. Predicted impact of COVID-19 on neglected tropical disease programs and the opportunity for innovation. Clinical Infectious Diseases. 2021;72(8):1463–1466.

31. Brooker SJ, Ziumbe K, Negussu N, Crowley S, Hammami M. Neglected tropical disease control in a world with COVID-19: an opportunity and a necessity for innovation. Transactions of The Royal Society of Tropical Medicine and Hygiene. 2021;115(3):205–207.

32. Borlase A, Blumberg S, Callahan EK, et al. Modelling trachoma post-2020: opportunities for mitigating the impact of COVID-19 and accelerating progress towards elimination. Transactions of the Royal Society of Tropical Medicine and Hygiene. 2021;115(3):213–221.

33. Ehrenberg JP, Utzinger J, Fontes G, et al. Efforts to mitigate the economic impact of the COVID-19 pandemic: potential entry points for neglected tropical diseases. Infectious diseases of poverty. 2021;10(01):4–13.

34. Khatri G, Mir SL, Hasan MM. Outbreak of monkeypox in south east asia; spotlight on bangladesh, pakistan and india. Annals of Medicine and Surgery. 2022;82:104361.

35. Ebola Outbreak Over in Uganda. 2023. https://www.cdc.gov/media/releases/2023/s0111-ebola-outbreak.html

36. Burki T. Cholera re-emerges in Haiti. The Lancet Infectious Diseases. 2023;23(3):288–289.

37. Mashal MT, Eltayeb D, Higgins-Steele A, et al. Partnership and domestic resource mobilization in Sudan for cVDPV2 outbreak response amidst multiple emergencies in 2020-2021. 2022;

38. McKimm-Breschkin JL, Hay AJ, Cao B, et al. COVID-19, Influenza and RSV: Surveillance-informed prevention and treatment–Meeting report from an isirv-WHO virtual conference. Antiviral Research. 2022;197:105227.

